# Assessment of Vaccination and Underreporting on COVID-19 Infections in Turkey Based On Effective Reproduction Number

**DOI:** 10.1101/2021.02.04.21251176

**Authors:** Tuğba Akman Yıldız, Emek Köse, Necibe Tuncer

## Abstract

In this paper, we introduce a SEIR type COVID-19 model where the infected class is further divided into subclasses with individuals in intensive care (ICUs) and ventilation units. The model is validated with the symptomatic COVID-19 cases, deaths, and the number of patients in ICUs and ventilation units as reported by Republic of Turkey, Ministry of Health for the period March 11, 2020 through May 30, 2020 when the nationwide lockdown is in order. COVID-19 interventions in Turkey are incorporated into the model to detect the future trend of the outbreak accurately. We tested the effect of underreporting and we found that the peaks of the disease differ significantly depending on the rate of underreporting, however, the timing of the peaks remains constant.

The lockdown is lifted on June 1, and the model is modified to include a time dependent transmission rate which is linked to the effective reproduction number ℛ_*t*_ through basic reproduction number ℛ_0_. The modified model captures the changing dynamics and peaks of the outbreak successfully. With the onset of vaccination on 13 January 2021, we augment the model with the vaccination class to investigate the impact of vaccination rate and efficacy. We observe that vaccination rate is a more critical parameter than the vaccine efficacy to eliminate the disease successfully.

## 1. Introduction

The novel coronavirus SARS-CoV-2 was firstly identified in Wuhan, China in December 2019 [1] and World Health Organization (WHO) characterised the situation as a pandemic in March, 2020 [2]. More than 92 million cases and 2 million deaths were reported worldwide (as of 17 January, 2020).

To asses the outbreak in Turkey in terms of precautions and interventions, first “COVID-19 Risk Assessment” was made public by Scientific Advisory Board for COVID-19 on January 22, 2020 and the first version of the booklet for guidelines was published on January 24, 2020 [3]. The first COVID-19 case was diagnosed on March 9, 2020 in Turkey, and the first fatality was reported on March 17, 2020 [4]. People showing symptoms of novel coronavirus infection have been monitored and contact tracing has been done. In addition, schools and universities moved to distance learning on March 16, 2020, some travel and other restrictions have been put into action to prevent the spread of the disease in public places and large cities of Turkey [4].

SARS-CoV-2 infection is quite different from previously known infectious diseases due to the asymptomatic cases reported in different parts of the world. For example, of all passengers and crew tested on Diamond Princess cruise ship, 58% of infected people did not show any symptoms [5], more strikingly of 408 people living in shelters in Boston, PCR-testing confirmed that 88.5% showed no symptoms [6]. For a review of asymptomatic case studies, we refer the reader to the study [7].

Unknown characteristics of SARS-CoV-2 infection led scientists to construct mathematical models incorporating the data so that long-term dynamics of the outbreak could be understood and efficient intervention strategies could be implemented by government authorities. There are numerous promising papers modeling COVID-19 pandemic in the literature; but, we here just mention some of them. For example, a SEIR (susceptible-exposed-infected-recovered) model including symptomatic and presymptomatic compartments has been used with both COVID-19 case and mortality data to predict the evolution of the disease and decide possible intervention strategies in the USA. The authors find that 95% compliance with mask use is vital to slow down the deaths caused by SARV-CoV-2 infection [8]. Ibio et al. explain the spread of the disease in Nigeria in terms of a SEIR-type model where the infectious compartment is divided into three subclasses as symptomatically-infectious, asymptomatically-infectious and hospitalized [9]. Garba et al. propose a generalized SEIR type model for the outbreak in South Africa by including the contaminated environment [10]. A SIDARTHE (susceptible, infected, diagnosed, ailing, recognized, threatened, healed and extinct) model is proposed by Giordano et al. where the difference between diagnosed and undiagnosed infectious individuals is included in the model and importance of social-distancing together with contact tracing and testing is underlined [11]. An age-dependent model is presented to determine the epidemic peak by using data of South Africa, Turkey and Brazil via data-fitting [12]. On the other hand, a SEIR type model is constructed for Turkey and a simulator called TURKSAS has been developed where authors predict the number of infected individuals to be approximately 123, 000 in March 21, 2020 [13]. With the use of Turkey data, a SEIR type model with quarantined compartment is developed to predict the peak with different intervention strategies [14].

Mathematical models can be used to discuss the influence of different intervention strategies so that appropriate precautions can be implemented before any epidemic peaks. For example, Iboi et al. show that use of face mask is very effective to control spread; but, it may not be possible due to economic limitations; so, other intervention strategies such as social-distancing and lockdown have to be implemented. [9] On the other hand, a novel *θ*-SEIHRD (with susceptible, exposed, infectious, undetected infectious, hospitalized, hospitalized that will die, dead, recovered, undetected recovered compartments) model is introduced for data of China and undetected infectious compartment is included to investigate the effect of this class to spread of the disease [15]. Ngonghala et al. propose a model including the isolated and quarantined compartments together with patients staying in intensive care unit (ICU) with US and New-York data [16]. They find that quarantine is the most effective intervention strategy among quarantine, tracing and face masks, to slow down the spread. Moreover, Eikenberry et al. compare different face masks efficacy and they find out that face mask use must be applied nation-wide even if a low quality mask is used [17], whereas Ngonghala et al. study the post-lockdown interventions in some states of the USA and find timing of lockdown and detection of pre-symptomatic and asymptomatically-infectious individuals is quite critical, together with face mask use [18]. Kennedy et al. develop a generalized SEIR model including a compartment for critical patients and they discuss the contribution of personal protection measures and social distancing to the number of patients in ICU [19]. They deduce that “a stepping-down approach every 80 days” can be applied to eliminate the second peak and to reduce the number of patients at ICU.

There is no special medicine that is known to treat SARS-CoV-2 infected people; but, new promising vaccines are still being developed to prevent people from infection [20]. In terms of vaccination models for COVID-19 pandemic, the number of studies is quite limited compared to modeling of non-pharmacologic interventions. For example, Gumel et al. present a model by decomposing susceptible compartment as vaccinated and unvaccinated subclasses by discussing stability and threshold for herd immunity [21]. In another study [22], a heterogeneous model based on the face mask use is presented and success of vaccination together with social-distancing is discussed. Another recent work takes 16 age groups into consideration and use of vaccination is optimized with four different objective functions [23]. On the other hand, a hypothetical case study for South Africa has been published and Mukandavire et al. suggest that social distancing is a key intervention strategy until an efficient vaccine is applied in the community [24].

Spread of a disease could be different from country to country and parameter estimation enables us to mimic real world phenomenon with the available data. As underlined in the study [25], it is crucial to determine the values of key parameters correctly, to update the model appropriately when new evidence becomes available and to understand the heterogeneity of the population in terms of the spread of the disease. Even though we can determine the contact rate via data-fitting, its constant value may cause simulation results to deteriorate, since transmission of the disease might change due to the update of government policies. Thus, time-dependent contact rate is preferable in some studies. For example, authors use the transmission rate as a slow-decaying continuous function inversely correlated with the parameter denoting the effects of the interventions [10]. In other studies [17, 26], time dependent transmission rate depending on minimum contact rate and the rate at which contact decreases is used. It is known that the reproduction number ℛ_0_ is a measure for the average number of secondary infections per infective [27]. However, it would be more flexible and accurate to use a time dependent measure computed from the daily reported cases, namely effective reproduction number ℛ_*t*_ or *R*(*t*), to catch the dynamics of the disease [28, 29, 30]. Indeed, the transmission rate can be written in terms of the effective reproduction number as a time dependent function in some SIR models [31, 32, 33].

In this study, we consider the COVID-19 outbreak in Turkey and develop a model using the data reported by Republic of Turkey, Ministry of Health [34] and incorporating the interventions implemented in Turkey [4]. We divide this study into some parts depending on the interventions put in place in Turkey to model the outbreak accurately. Therefore, we first construct a model with the compartments of individuals who are susceptible, exposed, asymptomatically infected, mildly infected, patients staying in ICUs and ventilation units, and we apply data-fitting from March 11, 2020 to May 31, 2020 with the data of symptomatic daily incidences, deaths, ICU prevalence and ventilator unit prevalence. Fitted results agree well with the reported data and global sensitivity analysis via PRCC shows that the most influential parameters leading to an increase in the number of patients in ICUs and ventilation units are recovery rates, whereas the contact rate associated with the asymptomatically infectious individuals and quarantine rate play an important role for the sensitivity of the cumulative number of cases. In addition, it is revealed that the infected individuals must be treated without letting the patient’s health deteriorate so that ICU and ventilation capacities will not be exceeded. After re-opening of public places with other updated interventions in Turkey on June 1, 2020 [35], we modify the model by computing a time dependent contact rate associated with the effective reproduction number from June 1, 2020 to January 3, 2021 in order to express the changing dynamics of the outbreak until January 3, 2021. On the other hand, we investigate the effect of underreporting which is a main concern in outbreaks due to unknown characteristics of the novel virus and prevalence of asymptomatic cases. Then, we compute the incidence rate under different underreporting scenarios. Until the beginning of the vaccination program, we perform model simulations with the constant contact rate associated with the basic reproduction number on January 3, 2021 (which is small due to interventions). Vaccination began on January 13, 2021 in Turkey [36], so we add a vaccinated compartment into our model to investigate the role of the rate of vaccination and efficacy of vaccines to eliminate the disease in Turkey. Our model successfully expresses the outbreak from March 11, 2020 to January 3, 2021 in Turkey and it can provide guidance to decide efficient intervention strategies before occurrence of future peak(s). The rest of the paper is organized as follows. Sec. 2 is devoted to the development of the first model, parameter estimation, derivation of basic and effective reproduction numbers. In Sec. 3, simulation results are presented with the fitted data and the global sensitivity analysis is discussed, and the effect of underreporting to incidence rate is investigated. Then, we present the model with vaccination compartment. The paper ends with summary and conclusion.

## 2. Mathematical model

Turkey’s population at time *t* is split into mutually exclusive seven compartments of individuals who are susceptible *S* := *S*(*t*), exposed *E* := *E*(*t*), asymptomatically infectious *I*_*A*_ := *I*_*A*_(*t*), mildly infectious (mildly infected symptomatic individuals including the ones staying in a health care facility but not in ICU) *I*_1_ := *I*_1_(*t*), patients staying in ICUs, *I*_2_ := *I*_2_(*t*), ventilation units *I*_3_ := *I*_3_(*t*) and recovered (now immune) *R* := *R*(*t*). Then, total population *N* := *N* (*t*) is written as

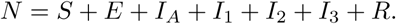

### Assumptions

- Some of the individuals in the class *I*_1_ can recover without staying at a health care facility.
- Only deaths due to SARS-CoV-2 are taken into account. We neglect deaths due to other diseases.
- Recovered individuals are not infectious.

The population of susceptible individuals decreases in size due to the interaction with the asymptomatically and symptomatically infectious individuals at the rates of *β*_*A*_ and *β*_1_, respectively. Interaction between susceptible and infectious individuals is reflected to the exposed class and the exposed class decreases in size at the rate of *p*_0_. Asymptomatically infectious individuals increase in size at the rate of (1 − *ρ*)*p*_0_*E* and they recover at the rate of *γ*_*A*_. The number of mildly infected individuals increases at the rate of *ρp*_0_ and this compartment decreases in size due to death, recovery or progression to the ICU (namely the class *I*_2_) at the rates of *µ*_1_, *γ*_1_, *p*_1_, respectively.

The number of patients in ICU increases as a result of the life-threatening symptoms of the disease at the rate *p*_1_ and this sub-population decreases due to death, recovery or the need of ventilation at the rates of *µ*_2_, *γ*_2_, *p*_2_, respectively. The compartment *I*_3_ increases in size due to the transfer of the patients in ICU to ventilated class at the rate of *p*_2_, while this class decreases in size due to death or recovery at the rates of *µ*_1_, *γ*_1_, respectively. The compartment *R* increases as a results of the successful recovery or treatment of the patients in the classes *I*_*A*_, *I*_1_, *I*_2_ and *I*_3_ at the rates of *γ*_*A*_, *γ*_1_, *γ*_2_, *γ*_3_, respectively. Then, we propose the following deterministic nonlinear system of ODEs to model the SARS-CoV-2 transmission in Turkey:

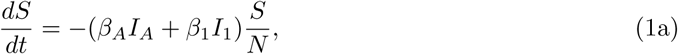

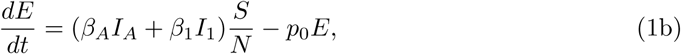

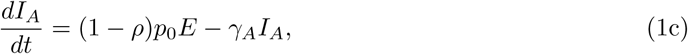

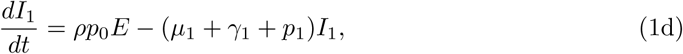

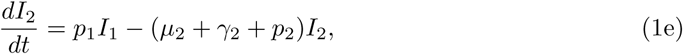

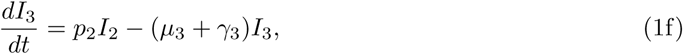

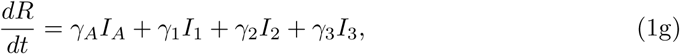

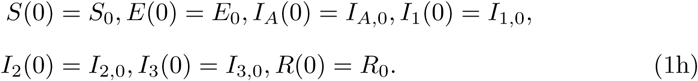

### 2.1. Parameter estimation

In this study, the data were taken from the Pandemic Monitoring Screen TURCOVID19 [34] where the authors explain the details of the platform in the letter to the editor [37]. Authors of this platform note that they collect up to date data from the daily announcements of Republic of Turkey, Ministry of Health [38], World Health Organization [39] and European Centre for Disease Prevention and Control [40].

Republic of Turkey, Minister of Health reported on March 19, 2020 that there were 25.466 ICU beds for adults (i.e. *I*_2,*max*_ = 25.000) [41]. Due to new regulations in health system and opening of new hospitals [42], this number has been expected to increase. On the other hand, the approximate number of ventilators was 17.000 on March 2020 (i.e. *I*_3,*max*_ = 17.000) [43]; but, domestic production of ventilators by different companies has led to an increase in the number of these devices since then [44].

We estimate the parameters of the model (1) from the daily reported symptomatic COVID-19 cases, deaths, ICU and ventilated patient prevalence in Turkey. For simplicity, let’s represent the model (1) in the following compact form:

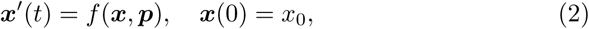

where ***x*** denotes the state variables and ***p*** denotes the parameters of the model of (1). What we observe, such as COVID-19 cases or deaths in Turkey, is a function of the state variables and parameters. To be specific, let ***y***(*t*) denote the observations, then

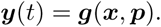

Here, we use daily symptomatic COVID-19 incidences and deaths in Turkey together with the daily number of patients in ICU and ventilation units at hospitals in Turkey. Parameters in the model cannot be determined from the literature due to ongoing discoveries of coronavirus and daily reported data for COVID-19 pandemic, but instead, obtained by data-fitting.

Let’s denote the symptomatic daily incidences as *y*_1_(*t*), deaths as *y*_2_(*t*), ICU prevalence as *y*_3_(*t*) and ventilator unit prevalence as *y*_4_(*t*), then in terms of state variables they have the following forms;

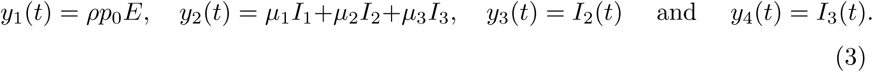

Similarly, let 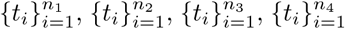 denote the discrete time when the number of symptomatic Corona cases, deaths, ICU and ventilation prevalence are measured, respectively. The measurement are contaminated with noise and thus,

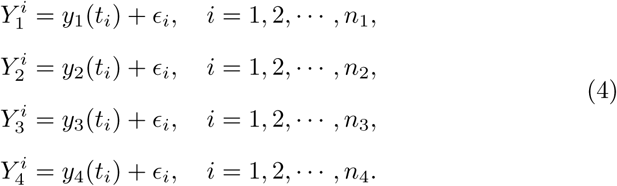

We estimated the parameters of the model (1) by solving the following optimization problem.

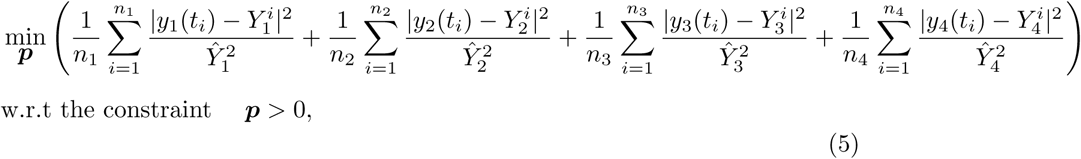

where *γ*_*A*,1,2,3_ *∈* (1*/*23, 1*/*4) [45, Table 3], *µ*_1,2,3_ *∈* (0, 1) [46] and *p*_0_ *∈* (1*/*14, 1*/*3) [47]. Here, *Ŷ*_1_, *Ŷ*_2_, *Ŷ*_3_ and *Ŷ*_4_ are the average daily symptomatic COVID-19 cases, deaths, ICU and ventilation unit prevalence, respectively, as shown below:

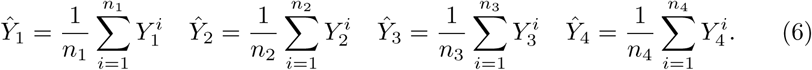

### 2.2. *Basic reproduction number* ℛ_0_

The model (1) has a disease-free equilibrium (DFE) point *ε*_0_ = (*S*_∗_, 0, 0, 0, 0, 0, *R*_∗_) where *N* (0) is the total initial population, *S*_∗_ := *N* (0) − *R*_∗_ and *R*_∗_ is a positive constant at the disease-free equilibria for *t ≥* 0 and 0 *≤ R*_∗_ *≤ N* (0). Stability of the DFE point is analyzed in terms of the next generation matrix [48, 49]. The matrices associated with the new infections ℱ and the transfer terms 𝒱 are found in two different ways by regarding *p*_0_*E* term as new infections or not, as explained in the study [48, p.36]. Then, it leads to alternative thresholds for our model as shown in Table 1 where 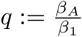.

**Table 1:** Decomposition of *f* leading to alternative computations of ℛ_0_.

|  | (a) | (b) |
| --- | --- | --- |
| $\mathcal{F}$ | $\begin{pmatrix} 0 \\ \beta_1(qI_A + I_1)\frac{S}{N} \\ 0 \\ 0 \\ 0 \\ 0 \\ 0 \end{pmatrix}$ | $\begin{pmatrix} 0 \\ \beta_1(qI_A + I_1)\frac{S}{N} \\ (1-\rho)p_0E \\ \rho p_0E \\ 0 \\ 0 \\ 0 \end{pmatrix}$ |
| $\mathcal{V}$ | $\begin{pmatrix} -\beta_1(qI_A + I_1)\frac{S}{N} \\ p_0E \\ -(1-\rho)p_0E + \gamma_A I_A \\ -\rho p_0E + (\mu_1 + \gamma_1 + p_1)I_1 \\ -p_1I_1 + (\mu_2 + \gamma_2 + p_2)I_2 \\ -p_2I_2 + (\mu_3 + \gamma_3)I_3 \\ -(\gamma_A I_A + \gamma_1 I_1 + \gamma_2 I_2 + \gamma_3 I_3) \end{pmatrix}$ | $\begin{pmatrix} -\beta_1(qI_A + I_1)\frac{S}{N} \\ p_0E \\ \gamma_A I_A \\ (\mu_1 + \gamma_1 + p_1)I_1 \\ -p_1I_1 + (\mu_2 + \gamma_2 + p_2)I_2 \\ -p_2I_2 + (\mu_3 + \gamma_3)I_3 \\ -(\gamma_A I_A + \gamma_1 I_1 + \gamma_2 I_2 + \gamma_3 I_3) \end{pmatrix}$ |
| $F$ | $\begin{pmatrix} 0 & q\beta_1\frac{S_0}{N(0)} & \beta_1\frac{S_0}{N(0)} \\ 0 & 0 & 0 \\ 0 & 0 & 0 \end{pmatrix}$ | $\begin{pmatrix} 0 & q\beta_1\frac{S_0}{N(0)} & \beta_1\frac{S_0}{N(0)} \\ (1-\rho)p_0 & 0 & 0 \\ \rho p_0 & 0 & 0 \end{pmatrix}$ |
| $V$ | $\begin{pmatrix} p_0 & 0 & 0 \\ -(1-\rho)p_0 & \gamma_A & 0 \\ -\rho p_0 & 0 & \mu_1 + \gamma_1 + p_1 \end{pmatrix}$ | $\begin{pmatrix} p_0 & 0 & 0 \\ 0 & \gamma_A & 0 \\ 0 & 0 & \mu_1 + \gamma_1 + p_1 \end{pmatrix}$ |
| $\mathcal{R}_0 = \rho(FV^{-1})$ | $\beta_1\left(q\frac{(1-\rho)}{\gamma_A} + \frac{\rho}{\mu_1 + \gamma_1 + p_1}\right)\frac{S_0}{N(0)}$ | $\sqrt{\beta_1\left(q\frac{(1-\rho)}{\gamma_A} + \frac{\rho}{\mu_1 + \gamma_1 + p_1}\right)\frac{S_0}{N(0)}}.$ |

### 2.3. Effective reproduction number ℛ_t_

Characterizing the infectivity of a pathogen through a population during an epidemic is important in terms of developing interventions. The basic reproduction number, ℛ_0_, which is the average number of secondary infections per infective, is used for this purpose. While ℛ_0_ is time and situation specific, ℛ_0_ *>* 1 implies an epidemic and the magnitude of ℛ_0_ is an indication of the transmissivity of the disease [27]. In the initial stages of an epidemic such as the COVID-19, however, the information that can be gleaned from ℛ_0_ decreases in time as its time-dependent version, the effective reproduction number, ℛ_*t*_, is more effective in reflecting the compliance with the non-pharmaceutical interventions and takes the changing number of susceptibles into account. Monitoring the ℛ_*t*_, therefore is more informative for managing an epidemic and for developing strategies for intervention. Estimating ℛ_*t*_ is a complicated statistical process and there are several commonly used methods to do so [28, 50, 51, 31, 27]. The two most popular methods are those of Cori et al. and Bettencourt and Ribiero [27, 28]. In [29], Gostic et al. caution against Bettencourt and Ribiero’s method as it requires structural assumptions about the epidemic and tends to underestimate the ℛ_*t*_ values and recommend Cori et al.’s method [27].

In our model, we incorporated the daily ℛ_*t*_ values in order to account for the crowd behavior, such as using face coverings, social distancing or sheltering in place. Although this is not very common in mathematical models of epidemiology, Buckman et al [31], Kiamari et al [32] and Linka et al [33] utilize the effective reproduction number ℛ_*t*_ in their compartmental SIR models as

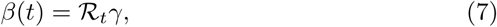

where *β* is the contact rate and 1*/γ* is the infectious period [31, 33].

Our calculation for the basic reproduction number ℛ_0_ reveals a relationship between ℛ_0_ and *β*_1_, *ρ, q, µ*_1_, *p*_1_, *γ*_1_ as given in Table 1, which allows us to write the transmission rate *β*_1_ in a time-dependent manner as *β*_1_(*t*), in terms of ℛ_*t*_ and the other parameters. Using the ℛ_0_ formulation in Table 1, we first set that the transmission rate *β*_1_(*t*) and 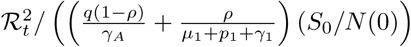 are directly proportional, we then experimentally found that the best fitting proportionality constant is 0.89. Note that there are two formulations of ℛ_0_, as explained in Table 1 and the ℛ_0_ formulation with the square root term provides a better fit to the data. Thus we set *β*_1_(*t*) as

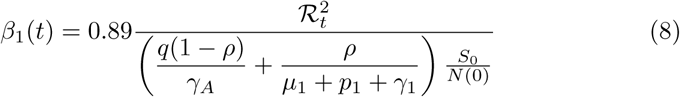

Because quarantine ceased to exist in Turkey on May 31, 2020 [35], we let *β*_1_(*t*) be updated daily henceforth based on our estimate of ℛ_*t*_ values. In other words, between June 1, 2020 and January 3, 2021 our model has a time-dependent transmission parameter. Note that, even though parameters were fit only using the data before June 1, 2020, simulation results agree very well with the data until January 2, 2021, end of the simulation (see Fig. 2).

We use the `R` package `EpiEstim`, written by Cori et al. [27] to estimate the daily ℛ_*t*_ values, based on a mean serial interval of 7 and standard deviation of 3.5 days. The estimate takes daily new case numbers as inputs, in addition to the mean and standard deviation of serial interval. The plot of ℛ_*t*_ values and the time-dependent *β*_1_(*t*) can be seen in Fig. 1.

**Figure 1:**
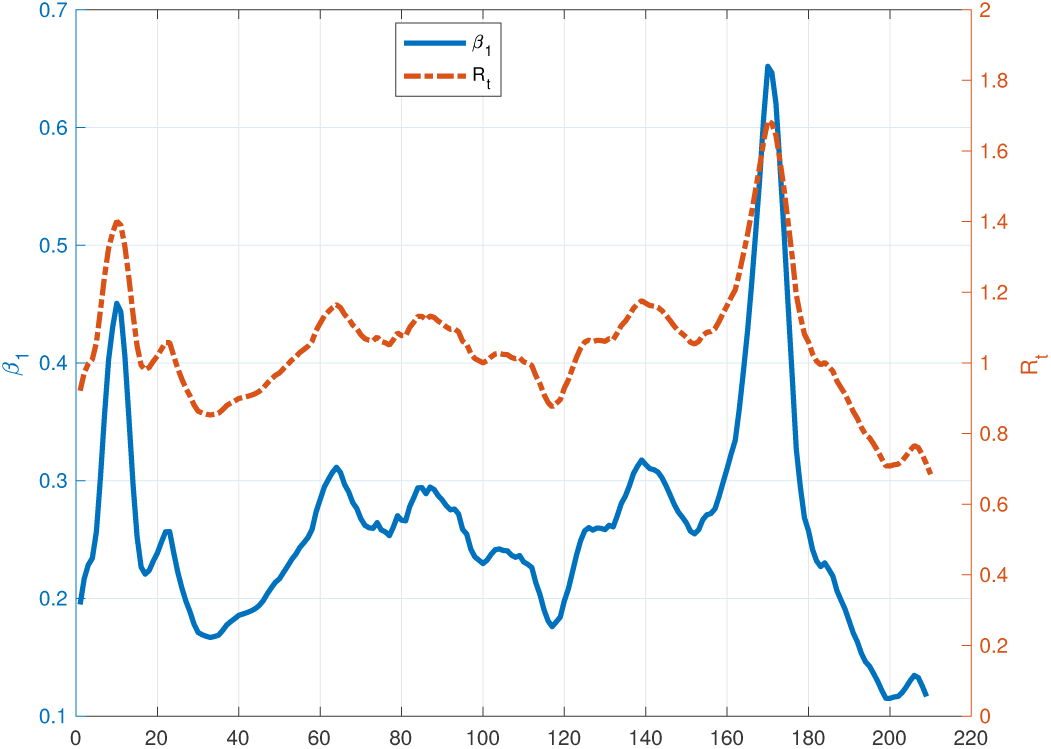
Contact rate *β*_1_(*t*) and effective reproduction number ℛ_*t*_ computed from the data on June 1st - January 3rd.

After solving the model (1) from March 11th to May 31st, we continue to simulate the model (9) from June 1st to January 13th with a time-dependent contact rate *β*_1_ as

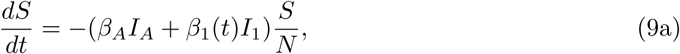

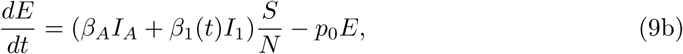

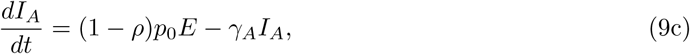

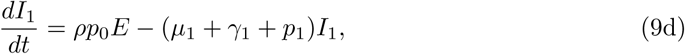

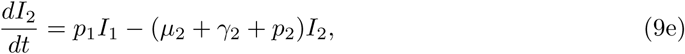

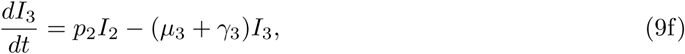

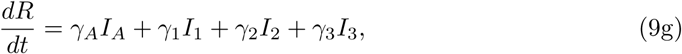

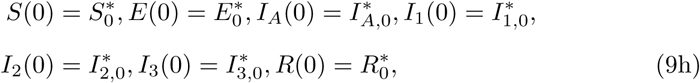

where the initial conditions 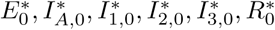 are the values of those variables on May 31st, 2020 obtained from the model (1). In addition, we fix the initial number of susceptible individuals on June 1st, 2020 approximately as 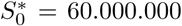, since quarantine restrictions were reorganized and public places were reopened on June 1st, 2020 [35].

Allowing the transmission rate depend on ℛ_*t*_ makes the model able to capture the change in regulations and social behaviors, which standard SEIR models would not be able to. The daily incidence, simulated by the model with time-dependent *β*_1_(*t*) is presented in Fig. 2.

We have shown that when the transmission rate depends on the effective reproduction number, the model fits to observed data better than if constant transmission rate is used. In order to predict the future of the epidemic, we consider three scenarios for how ℛ_*t*_ could perform, namely; 1) ℛ_*t*_ has an exponential increase of 0.5%, daily; 2) ℛ_*t*_ acts as a sawtooth function where it increases at a daily rate of 0.5% for 60 days and decreases at the same rate for 30 days; 3) ℛ_*t*_ behaves like a sine function, accounting for seasonal changes. Each of the three scenarios are run for 6 months after January 3rd, 2021 which marks the end of our ℛ_*t*_ calculations from data. The comparison of three ℛ_*t*_ time series and the simulations for the 3 scenarios for ℛ_*t*_ are presented below in Fig. 3.

**Figure 2:**
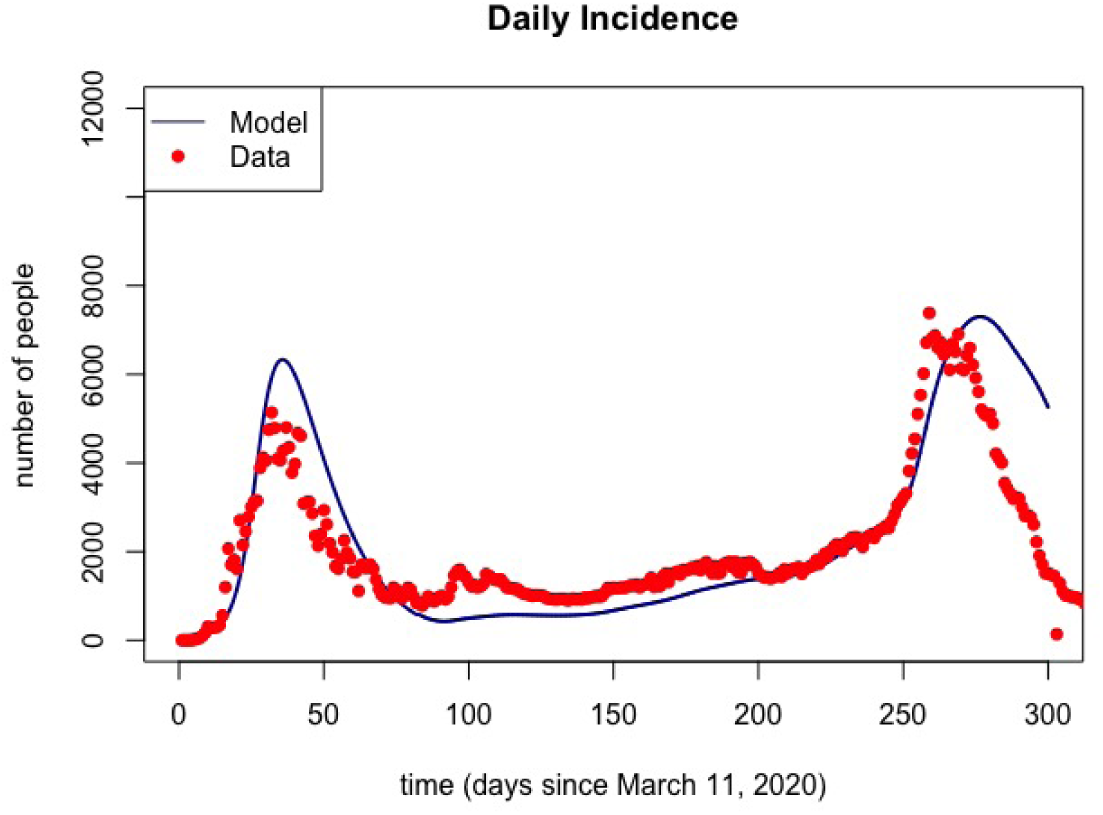
The incidence as simulated by the model with time-dependent transmission rate.

**Figure 3:**
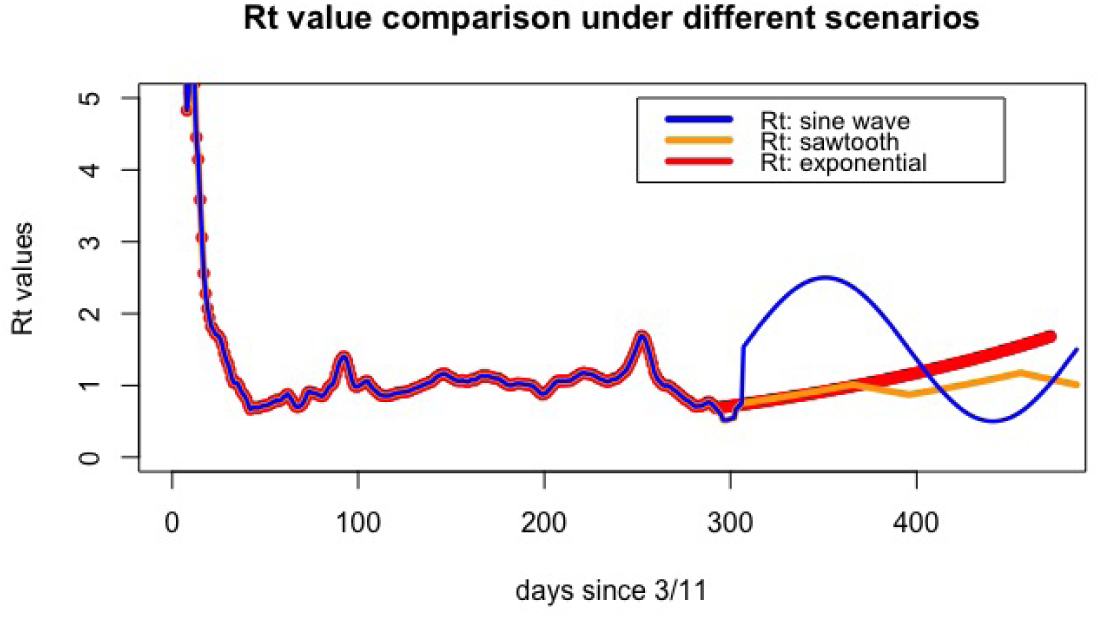
The three ℛ_*t*_ scenarios.

Although there are sizeable differences in the daily incidence numbers under the three ℛ_*t*_ scenarios (see Fig. 4), we have found comparable behavior for all ℛ_*t*_ scenarios in terms of the final wave of infections and their severity on the ICU and ventilator capacity. It is certain that as long as ℛ_*t*_ is allowed to remain mostly above 1, the healthcare system will be overwhelmed in the next wave of infections as shown in Fig. 5.

**Figure 4:**
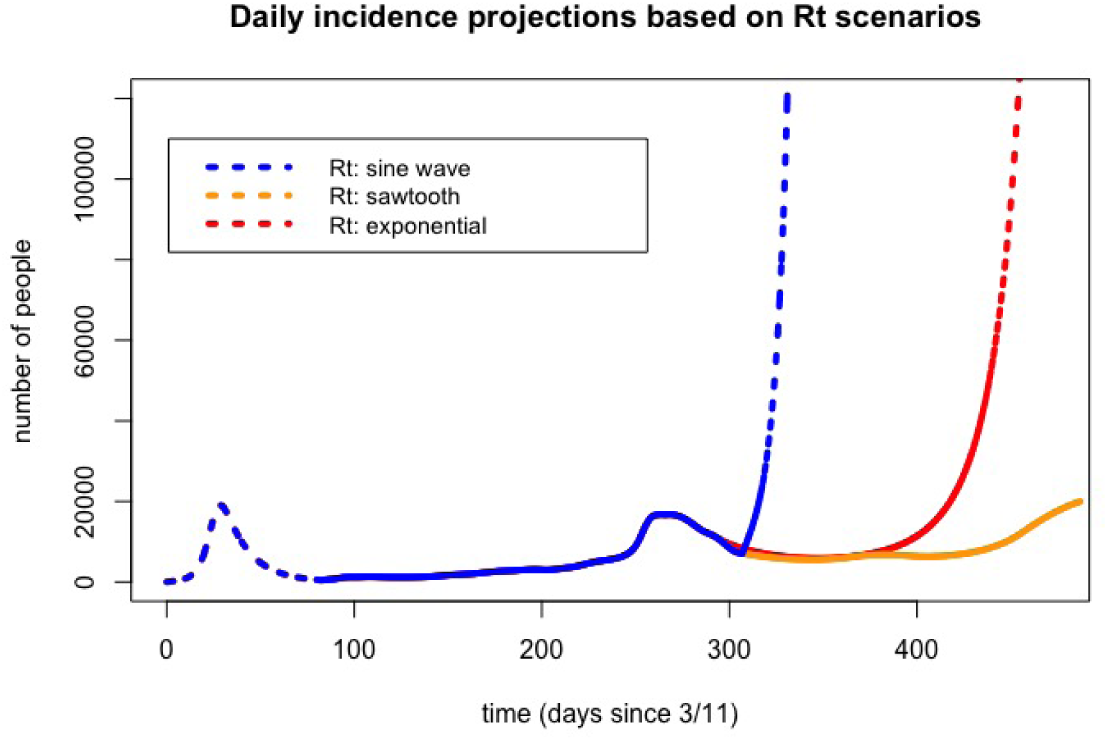
The daily incidence predictions under the three ℛ_*t*_ scenarios.

**Figure 5:**
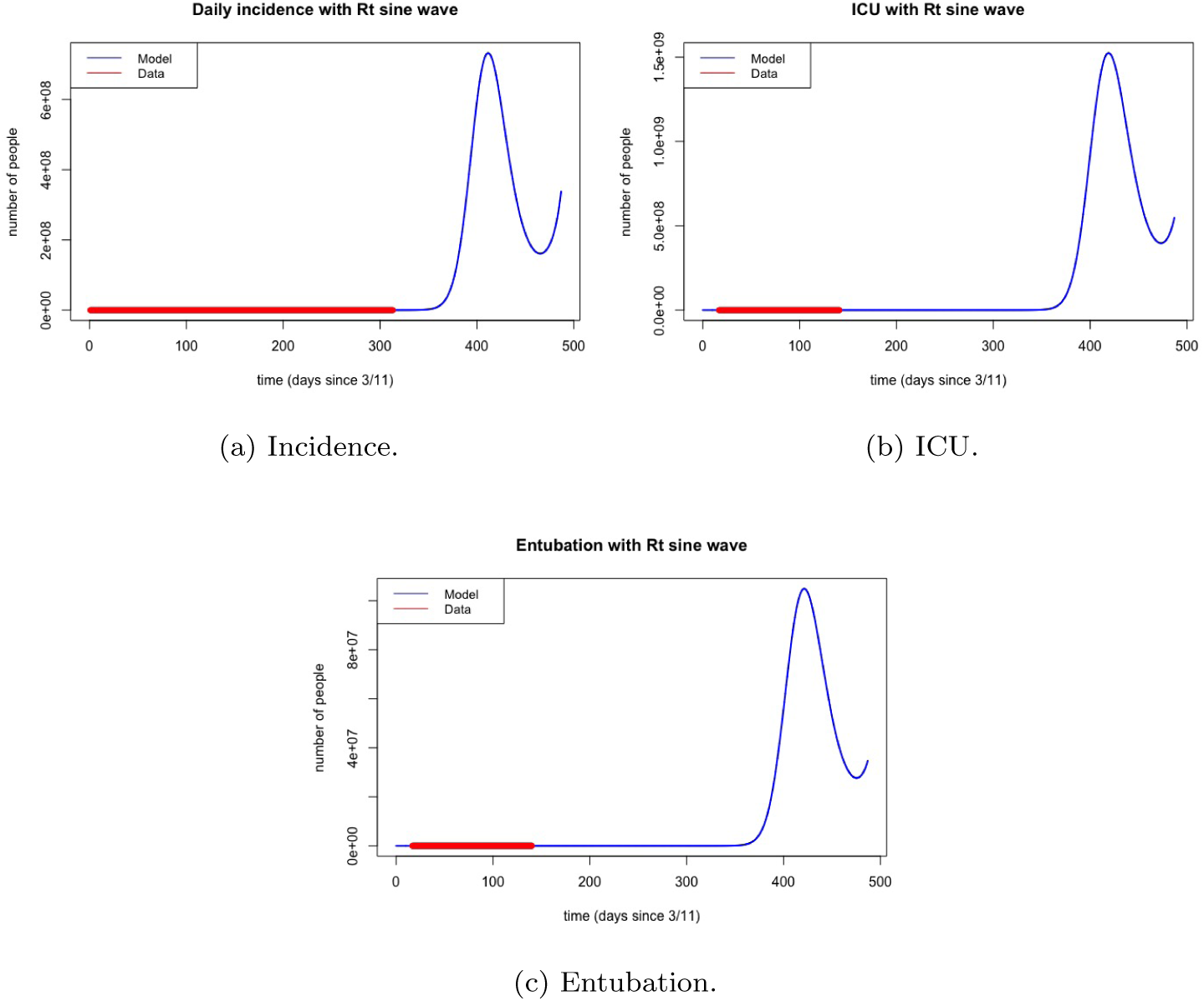
Simulation results for daily incidence, ICU and entubation under sine wave function for ℛ_*t*_, after January 3rd, 2021.

## 3. Simulation results

In this section, we begin with the results obtained from data fitting and global sensitivity analysis via PRCC, then proceed with the effects of underreporting of cases and vaccination strategies. We numerically solve the model (1) from March 11, 2020 to May 31, 2020 and fit the data [34]. The first symptomatic Corona case was tested positive on March 11, 2020; while the first death was recorded in March 17, 2020 in Turkey. The patients at ICU and ventilated individuals were reported first on March 27, 2020 [34].

Some intervention strategies such as quarantine of people older than 65 years as of March 21, 2020 and people younger than 20 years old as of April 3, 2020, closing of schools and universities as of 16 March 2020 have been incorporated into the model [4]. Since some portion of susceptible individuals have been isolated, they left the susceptible class at a rate of *c* due to these intervention strategies applied in Turkey. Then, it leads us to modify Eq. (1a) for data-fitting purposes as follows:

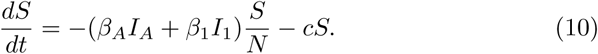

While applying data fitting for the parameters together with the initial conditions *E*_0_, *I*_*A*,0_, *I*_1,0_, *I*_2,0_ and *I*_3,0_, we set *S*_0_ = 80.000.000 as Turkey’s population is about 83.000.000 [52] and we find the following fitted initial conditions, which are different than the reported data:

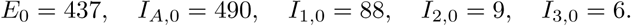

After inserting these initial conditions into the model, we find the values given in the Table 2 by data-fitting using `fminsearchbnd` function in MATLAB [53].

**Table 2:** Values of the parameters.

| Parameter | Description | Value ($\text{day}^{-1}$) |
| --- | --- | --- |
| $\gamma_A$ | Recovery rate for $I_1$ | 0.108457 |
| $\gamma_1$ | Recovery rate for $I_1$ | 0.0953836 |
| $\gamma_2$ | Recovery rate for $I_2$ | 0.231285 |
| $\gamma_3$ | Recovery rate for $I_3$ | 0.239209 |
| $p_1$ | Rate of progression from $I_1$ to $I_2$ | 0.0298359 |
| $p_2$ | Rate of progression from $I_2$ to $I_3$ | 0.138963 |
| $\mu_1$ | Death rate for $I_1$ | 8.09449e-06 |
| $\mu_2$ | Death rate for $I_2$ | 0.0605032 |
| $\mu_3$ | Death rate for $I_3$ | 0.000293138 |
| $\beta_A$ | Contact rate associated with $I_A$ | 1.01051 |
| $\beta_1$ | Contact rate associated with $I_1$ | 2.09096 |
| $p_0$ | Rate of progression from $E$ to $I_1$ | 0.0728926 |
| $\rho$ | Fraction of symptomatic infectious individuals | 0.28735 |
| $c$ | Quarantine rate for $S$ | 0.207711 |

### Remark 1

*With the parameter values in Table 2, the basic reproduction numbers* ℛ_0_ *in Table 1 are obtained as 11.4377 and 3.3820 with the formulas a and b, respectively*.

We present the simulation results for symptomatic Corona cases, deaths and patients in ICUs and ventilated units in Fig. 6 together with the reported data. We can see that data-fitting with four different classes gives good results and our model is capable of capturing the evolution of the disease in Turkey.

**Figure 6:**
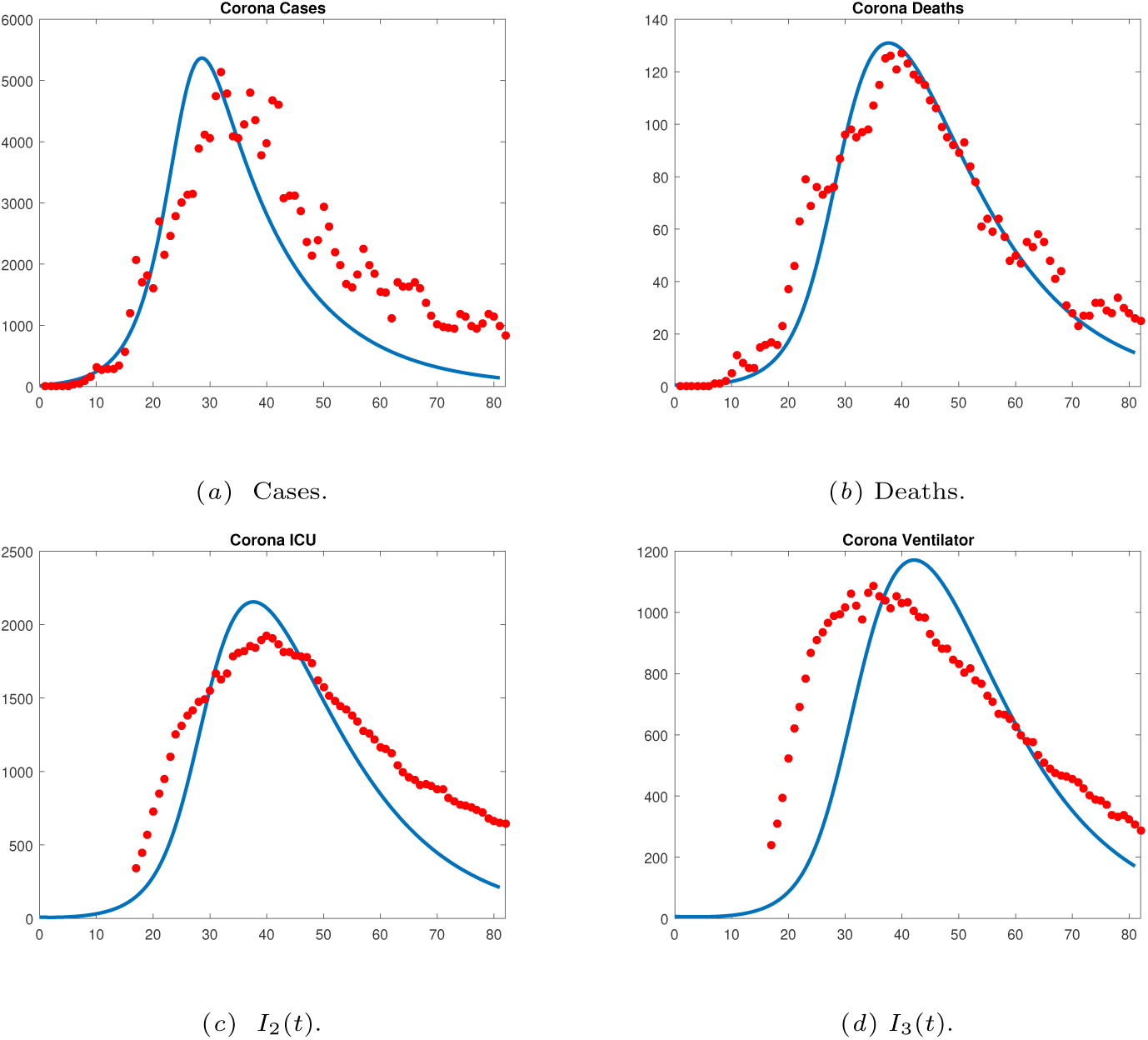
Model prediction versus data with fitted results.

### 3.1. Global sensitivity analysis via PRCC

We investigate the sensitivities of the compartments *I*_2_(*t*) and *I*_3_(*t*) and the cumulative number of cases to changes in the values of the parameters in the model (1) with Eq. 10 at days around the peak of the first wave (i.e. days 30, 37, 43 and 50). In order to do that, we perform global sensitivity analysis via the partial rank correlation coefficient (PRCC) [54] where it measures the influence of each parameter to a prespecified model variable. Indeed, PRCC assigns a value between -1 and +1 to each relationship and the magnitude of this quantity determines the strength whereas the sign reveals the trend of the relationship between the parameter and the variable.

To perform global sensitivity analysis, we construct a uniformly distributed sample space for each parameter with 1000 sample values. The intervals for each parameter other than *ρ* are chosen starting from half to twice the baseline parameter, where the interval for *ρ* is set from zero to one. For each time point, we depict the results in Fig. 7 and we can mention the most influential parameters to these groups. For example, there is an inverse relation between the compartment *I*_2_(*t*) and the parameters *γ*_1_, *γ*_2_ (recovery rates of the subgroups *I*_1_ and *I*_2_(*t*)), whereas the subgroup *I*_3_(*t*) is inversely sensitive to the parameters *γ*_1_ and *γ*_3_ (recovery rates of the subgroups *I*_1_ and *I*_3_(*t*)). On the other hand, as the fraction of symptomatic infectious individuals increases, the number of patients in ICUs is expected to increases; while the number of patients staying in ventilation unit will increase if the number of severe cases in ICUs cannot be cured successfully. In terms of the cumulative number of Corona cases, it is vital to keep the contact rate *β*_*A*_ under control and quarantine rate is the most influential parameter reducing the cumulative cases. As time passes, we can observe that these parameters stay as the most influential ones. Thus, these results reveal the importance of recovery rates to control the capacities of ICUs and ventilation units, and isolation or quarantine strategies play an important role to decrease the cumulative cases.

**Figure 7:**
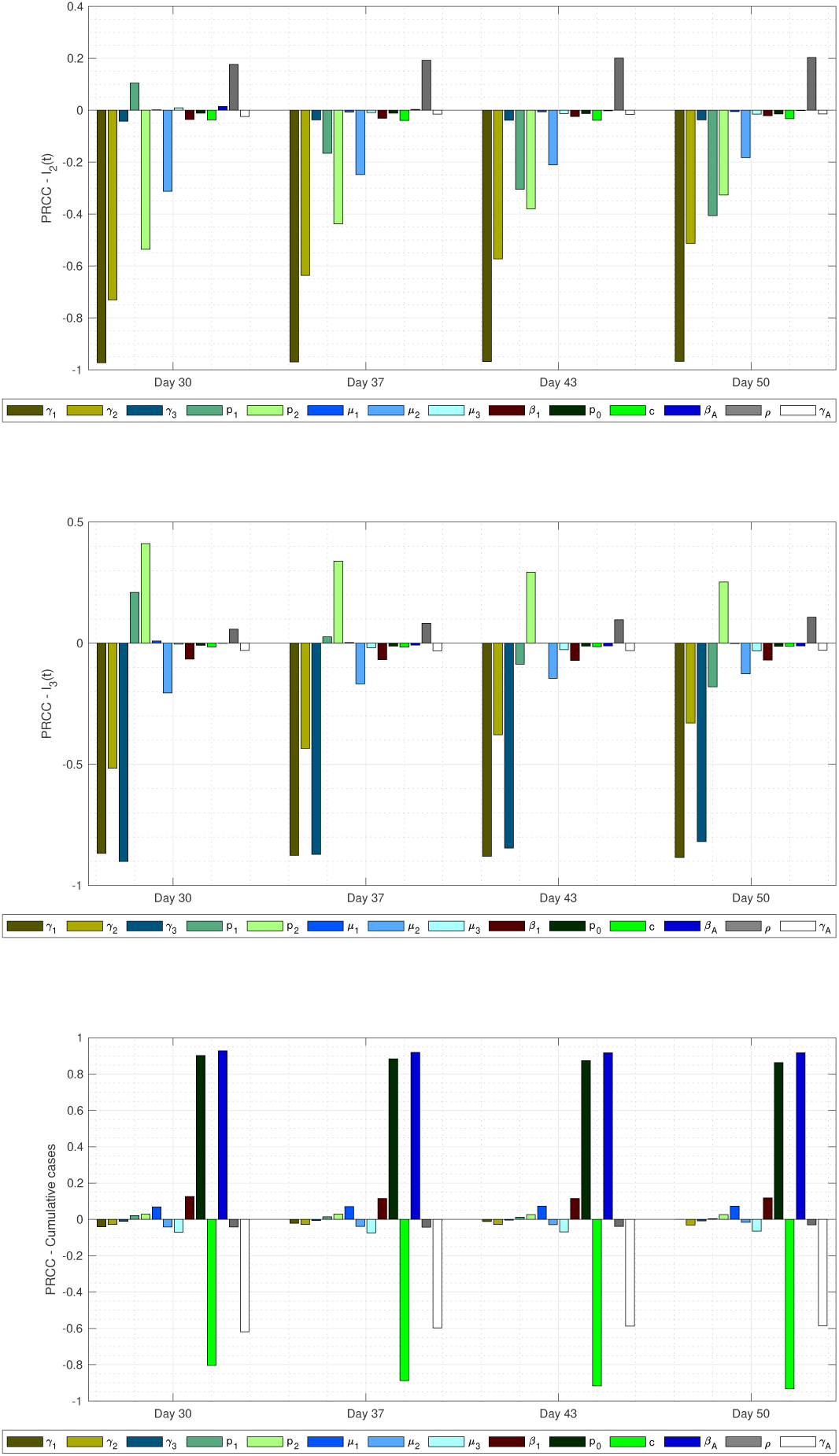
Sensitivity analysis for *I*_2_, *I*_3_ and cumulative cases.

### 3.2. Influence of underreporting

Due to the significant portion of all infections being asymptomatic and the insufficient testing capacity early in the pandemic, the number of cases of COVID-19 are severely underreported. CDC states that 6.7 in 7.7 symptomatic cases are unreported [55]. This number is expected to be higher for asymptomatic cases. Thus, underreporting is a concern from a public health perspective and it is important to understand its impact on the effective reproduction number, as well as the peaks of each wave in the epidemic. We have tested the effects of different underreporting rates on the incidences and have found that the peaks of the disease differ significantly depending on the rate of underreporting, as presented in Fig. 8, however, the timing of the peaks remains constant with or without underreporting. The effective reproduction numbers between the case counts with varying rates of underreporting do not differ significantly, also, as shown in Fig. 9. Since the number of people infected by a single infective is not influenced by the number of underreported cases, the difference in ℛ_*t*_ values calculated with and without underreporting is negligible.

**Figure 8:**
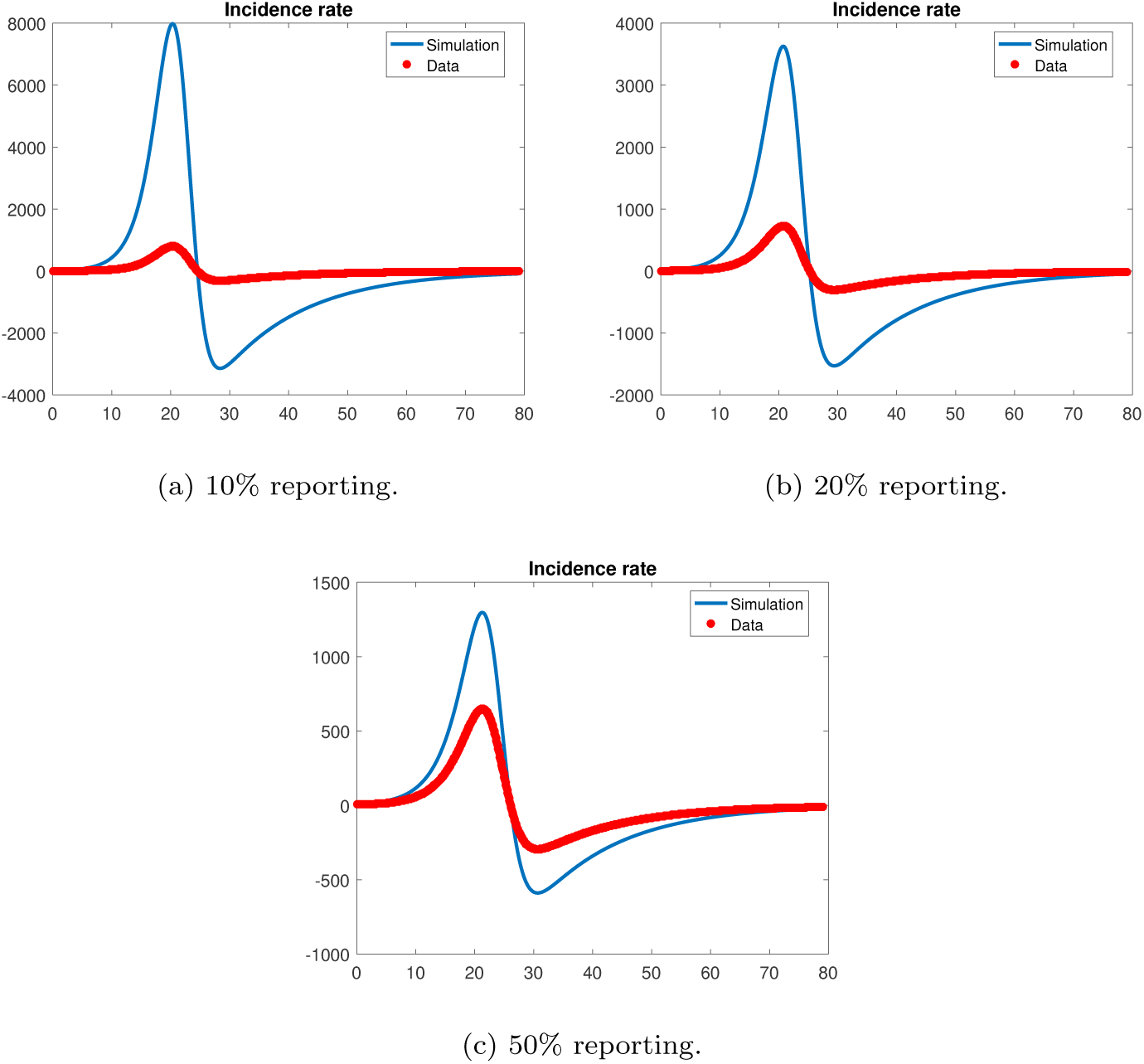
Effect of under-reporting to incidence rate.

**Figure 9:**
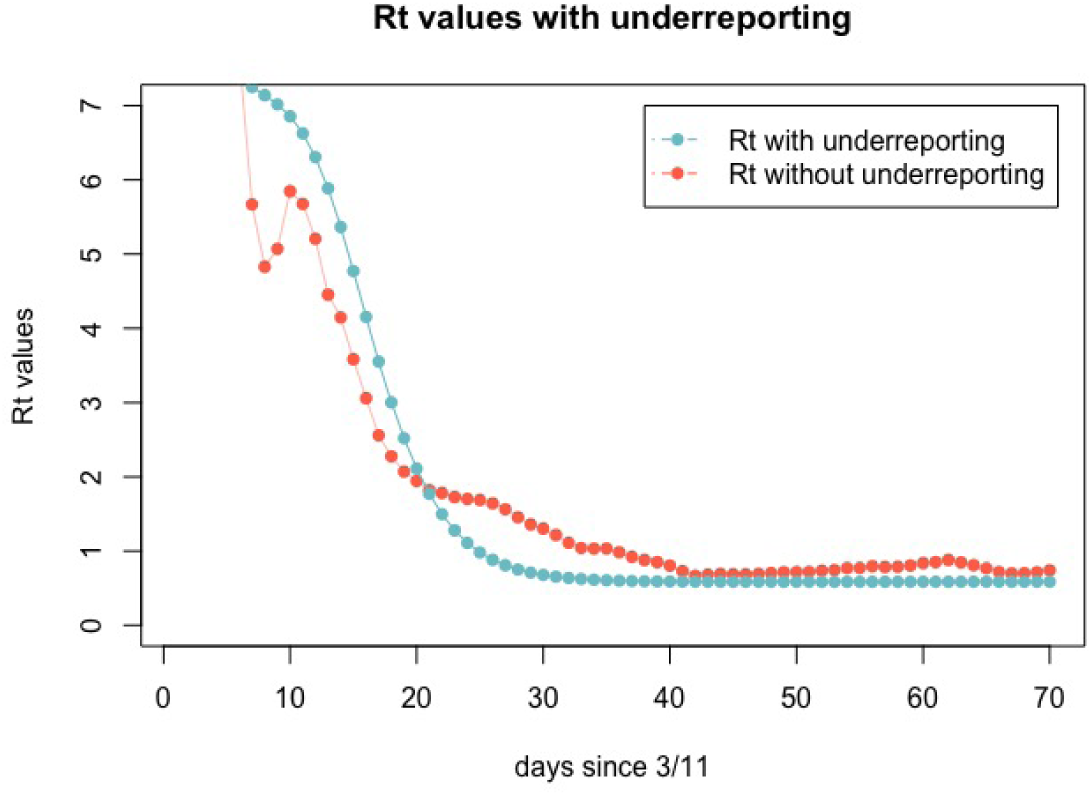
Comparison of ℛ_*t*_ estimates with and without underreporting.

### 3.3. Influence of vaccination

We incorporate the effective reproduction number ℛ_*t*_ to compute the time-dependent contact rate *β*_1_(*t*) between June 1, 2020 - January 3, 2021, as mentioned before. We continue to simulate the model (1) with the constant transmission rate 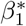 from January 4, 2021 to January 13, 2021 where 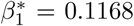 is the value of *β*_1_(*t*) computed from ℛ_*t*_ associated with the data on January 3, 2021. With the start of the vaccination program with Sinovac’s vaccine on January 13, 2021 in Turkey [36], we construct the extended model (11) with the vaccinated compartment as follows:

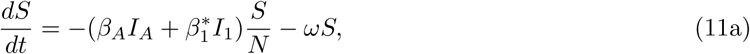

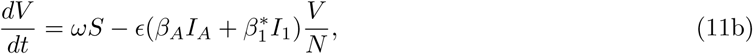

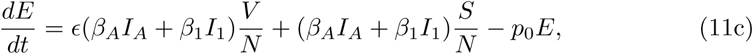

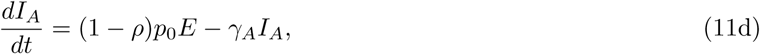

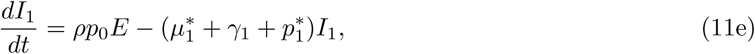

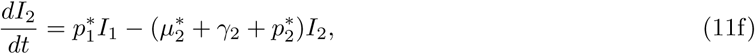

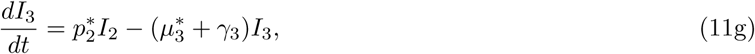

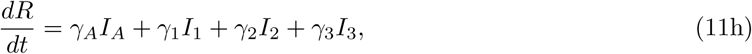

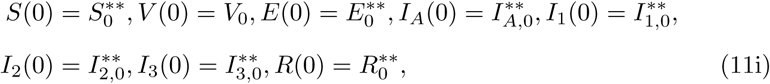

where *ω* is the rate at which susceptible individuals are vaccinated and *ϵ* is the reduction of infectiousness computed from efficacy of the vaccine. In addition, the parameter *p*^∗^ = *ηp* is defined for the parameters *p*_1_, *p*_2_, *µ*_1_, *µ*_2_, *µ*_3_ with 0 *< η <* 1, since vaccination decreases the number of infectious individuals and it is reflected to Corona deaths, the number of ICU patients and ventilated patients positively. To the best of our knowledge, there is no available information to determine *η*; so, we fix it as 0.5 to see the overall contribution of vaccination to the outbreak. Initial conditions are the values obtained from the solution of the model (1) on January 13, 2021 with the contact rate of 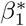. Values of the other parameters are the same as in Table 2, where *ω* and *ϵ* are changed to represent different vaccination rates and vaccine types.

We present the simulation results for deaths, daily cases and cumulative number of cases with and without vaccination from March 11, 2020 to September 13th, 2021 in Fig. 10. We fix the vaccination rate as *ω* = 0.001, which means that approximately 0.1% of the susceptible compartment is vaccinated per day, that is, approximately 60.000 individuals are vaccinated per day. Moreover, we observe the contribution of vaccination for eight months. In Fig. 10 on the left panel, the results are presented for vaccines which are 95 % effective against SARS-CoV-2 such as BNT162b2 (produced by Pfizer and BioNTech [56]) and mRNA-1273 (produced by Moderna [57]). On the right panel, results are presented for a vaccine showing an average 70% efficacy such as AZD1222 vaccine (produced by Astrazeneca and researchers from University of Oxford) [58]. We note that Sinovac’s vaccine was reported as approximately 91.25% effective in Turkey whereas it is 78% effective to mild to severe cases in Brazil; but its efficacy is calculated about 50% when applied to health workers in Brazil [59]. We present the results for *ϵ* = 0.3 and *ϵ* = 0.05 which correspond to 70% and 95% efficacies, respectively, since this range includes that of Sinovac’s vaccine.

**Figure 10:**
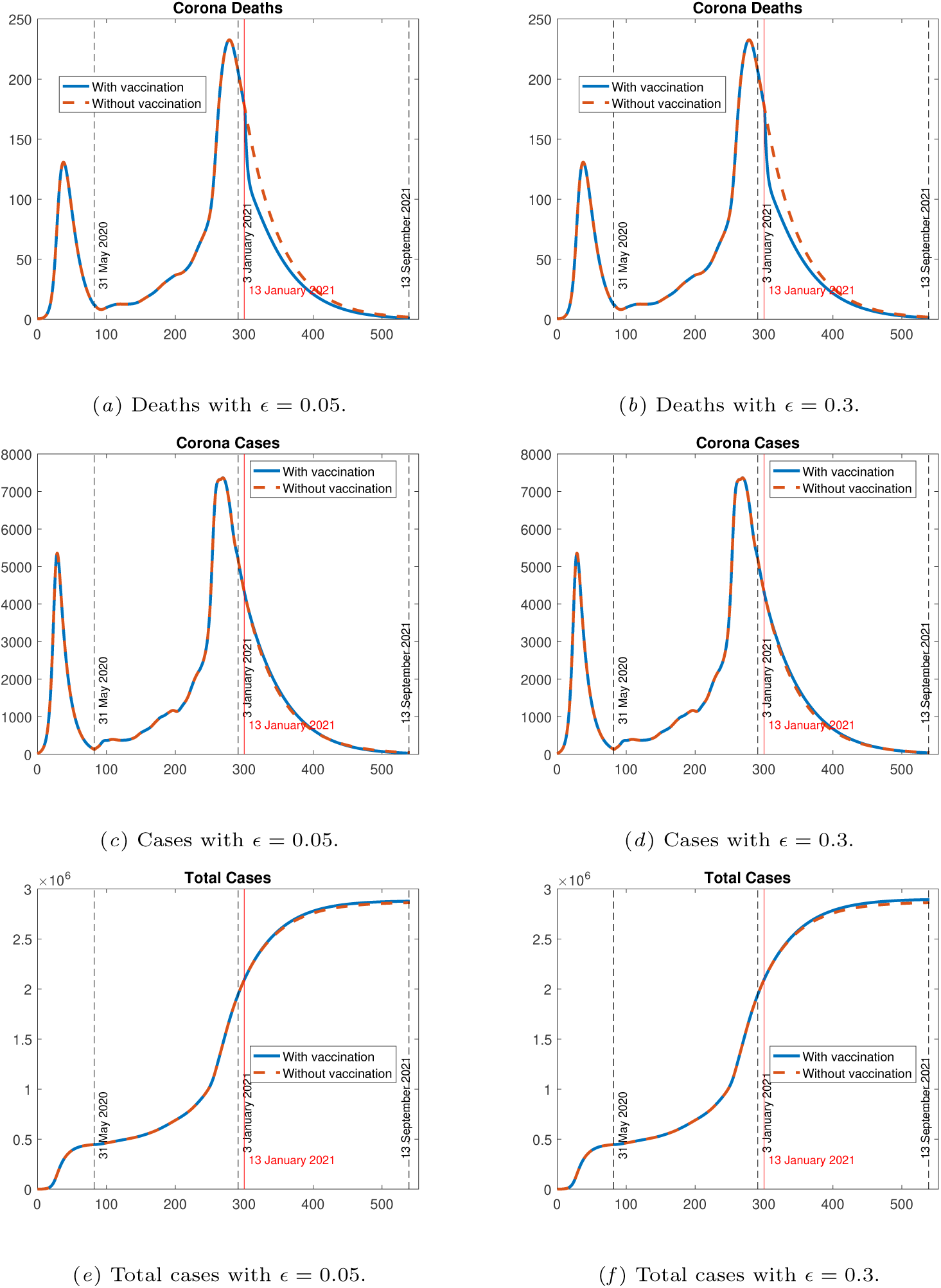
Simulation results w/o vaccination with *ω* = 0.001 and different values of *ϵ*.

We immediately observe from Fig. 10 that vaccination leads to a decrease in the number of deaths, daily cases and the cumulative number of cases. When we compare the left and right panels, we can see the contribution of vaccine. Even though the difference between the curves associated with vaccinated and unvaccinated cases is quite small, our model predicts that a vaccine with 95% efficacy is more successful than a vaccine with 30% efficacy, as expected. Therefore, we can say that vaccine’s efficacy is neglectable if vaccination rate and transmission rate are quite small.

We depict the results for Corona deaths, daily cases and cumulative number of cases with and without vaccination by fixing the efficacy as *ϵ* = 0.05 and using three different vaccination rates of *ω* as 0.01 (the left panel), 0.1 (the middle panel) and 0.9 (the right panel) in Fig. 11. As we increase the vaccination rate, curves associated with the number of deaths and Corona cases decrease faster whereas cumulative number of cases reaches to a smaller steady-state value. We conclude that vaccination rate is quite critical to eliminate the disease fast.

**Figure 11:**
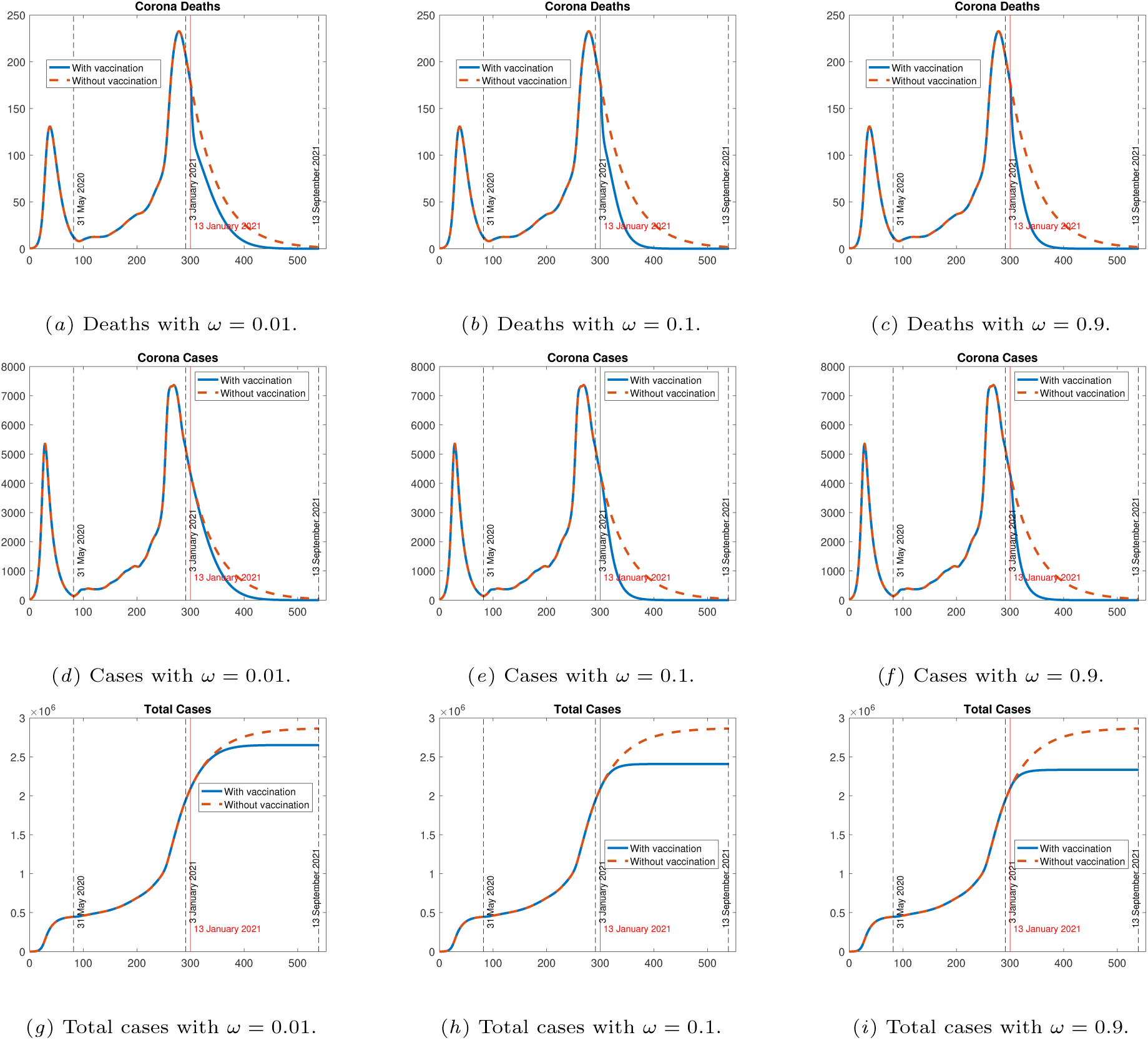
Simulation results w/wo vaccination with *ϵ* = 0.05 and different values of *ω*.

## 4. Summary and conclusion

Compartmental models are widely used in modeling epidemics and although they are very informative about the general behavior of the disease, they fail to capture the effects of non-pharmaceutical interventions or changes in crowd behavior. One way to mitigate this problem is to use time-dependent parameters. In this paper, we propose incorporating the effective reproduction number as a proxy for crowd behavior in terms of adhering to the changing interventions. The most natural way to include the ℛ_*t*_ is by describing the transmission parameter *β*_1_ as a function of ℛ_*t*_. Even though the parameters are estimated using 82 days of data, the model is capable of adjusting to the changes in the outbreak. In other words, our model provides an accurate and consistent fit to the observed case counts, including the multiple waves of infection and thus, enables us to experiment with the effects of vaccination in a reliable way. We have found that underreporting, which is a salient property of COVID-19 specifically, does not change the time of the peaks of each wave, but only the maximum incidence. On the other hand, we observe that rate of vaccination of susceptible individuals is a critical parameter to eliminate the disease even if a vaccine with a small efficacy like 70% is used.

Different age groups could be incorporated into the model to investigate the contribution of vaccination of prioritized age groups. Moreover, this model could be extended by taking waning immunity into consideration and this could be discussed in a future study as more information will be available related to the impact of vaccination.

## Data Availability

We have used data from TURCOVID19 project, which is presents publicly available data from Turkish Ministry of Health. The data can be found here: https://turcovid19.com/acikveri/

## CRediT authorship contribution statement

**Tuğba Akman Yıldız**: Conceptualization, Methodology, Software, Formal analysis, Investigation, Writing - Original Draft, Writing - Review & Editing, Supervision, Project administration. **Emek Köse**: Conceptualization, Methodology, Software, Formal analysis, Writing - Review & Editing. **Necibe Tuncer**: Conceptualization, Methodology, Formal analysis, Writing - Review & Editing.

## References

[1] R. Lu, X. Zhao, J. Li, P. Niu, B. Yang, H. Wu, W. Wang, H. Song, B. Huang, N. Zhu, et al., Genomic characterisation and epidemiology of 2019 novel coronavirus: implications for virus origins and receptor binding, The Lancet 395 (10224) (2020) 565–574. doi:10.1016/S0140-6736(20)30251-8.

[2] World health organization, WHO announces COVID-19 outbreak a pan-demic, Available: https://www.euro.who.int/en/health-topics/health-emergencies/coronavirus-covid-19/news/news/2020/3/who-announces-covid-19-outbreak-a-pandemic, (Accessed on 13.01.2021) (12 March 2020).

[3] Health system response monitor COVID-19, Policy responses for Turkey, Available: https://www.covid19healthsystem.org/countries/turkey/livinghit.aspx?Section=3.2%20Managing%20cases&Type=Section (Accessed on 13.01.2021) (November 2020).

[4] Y. Demirbilek, G. Pehlivantürk, Z. Ö. Ö zgüler, E. A. Meşe, COVID-19 outbreak control, example of ministry of health of Turkey, Turkish journal of medical sciences 50 (SI-1) (2020) 489–494. doi:10.3906/sag-2004-187.

[5] A. Sakurai, T. Sasaki, S. Kato, M. Hayashi, S.-i. Tsuzuki, T. Ishihara, M. Iwata, Z. Morise, Y. Doi, Natural history of asymptomatic SARS-CoV-2 infection, New England Journal of Medicine (2020). doi:10.1056/NEJMc2013020.

[6] T. P. Baggett, H. Keyes, N. Sporn, J. M. Gaeta, Prevalence of SARS-CoV-2 infection in residents of a large homeless shelter in Boston, Jama 323 (21) (2020) 2191–2192. doi:10.1001/jama.2020.6887.

[7] E. A. Meyerowitz, A. Richterman, I. I. Bogoch, N. Low, M. Cevik, Towards an accurate and systematic characterisation of persistently asymptomatic infection with SARS-CoV-2, The Lancet Infectious Diseases (2020). doi:10.1016/S1473-3099(20)30837-9.

[8] IHME COVID-19 Forecasting Team, Modeling COVID-19 scenarios for the United States,Nature medicine 27 (2021) 94–105. doi:10.1038/s41591-020-1132-9.

[9] E. A. Iboi, O. O. Sharomi, C. N. Ngonghala, A. B. Gumel, Mathematical modeling and analysis of COVID-19 pandemic i. Nigeria, Mathematical Biosciences and Engineering 17 (2020) 7192–7220. doi:10.3934/mbe.2020369.

[10] S. M. Garba, J. M.-S. Lubuma, B. Tsanou, Modeling the transmission dynamics of the COVID-19 Pandemic in Sout. Africa, Mathematical biosciences 328 (2020) 108441. doi:10.1016/j.mbs.2020.108441.

[11] G. Giordano, F. Blanchini, R. Bruno, P. Colaneri, A. Di Filippo, A. Di Matteo, M. Colaneri, Modelling the COVID-19 epidemic and implementation of population-wide interventions i. Italy, Nature Medicine (2020) 1– 6doi:10.1038/s41591-020-0883-7.

[12] S. Djilali, B. Ghanbari, Coronavirus pandemic: A predictive analysis of the peak outbreak epidemic in South Africa. Turkey, and Brazil, Chaos, Solitons & Fractals (2020) 109971 doi:10.1016/j.chaos.2020.109971.

[13] A. Ucar, S. Arslan, M. Y. Ozdemir, Nowcasting and forecasting the spread of COVID-19 and healthcare demand i. Turkey, A modelling study, medRxiv (2020). doi:10.1101/2020.04.13.20063305.

[14] M. Demir, M. M. Wise, S. Lenhart, et al., Modeling COVID-19: Forecasting and analyzing the dynamics of the outbreak in Hubei and Turkey, medRxiv (2020). doi:10.1101/2020.04.11.20061952.

[15] B. Ivorra, M. R. Ferrández, M. Vela-Pérez, A. Ramos, Mathematical modeling of the spread of the coronavirus disease 2019 (COVID-19) taking into account the undetected infections. the case o. China, Communications in nonlinear science and numerical simulation 88 (2020) 105303. doi:10.1016/j.cnsns.2020.105303.

[16] C. N. Ngonghala, E. Iboi, S. Eikenberry, M. Scotch, C. R. MacIntyre, M. H. Bonds, A. B. Gumel, Mathematical assessment of the impact of non-pharmaceutical interventions on curtailing the 2019 nove. Coronavirus, Mathematical Biosciences (2020) 108364.

[17] S. E. Eikenberry, M. Mancuso, E. Iboi, T. Phan, K. Eikenberry, Y. Kuang, E. Kostelich, A. B. Gumel, To mask or not to mask: Modeling the potential for face mask use by the general public to curtail the COVID-19 pandemic, Infectious Disease Modelling (2020).

[18] C. N. Ngonghala, E. A. Iboi, A. B. Gumel, Could masks curtail the postlockdown resurgence of COVID-19 in the us?, Mathematical biosciences 329 (2020) 108452. doi:10.1016/j.mbs.2020.108452.

[19] D. M. Kennedy, G. J. Zambrano, Y. Wang, O. P. Neto, Modeling the effects of intervention strategies on COVID-19 transmissio. dynamics, Journal of Clinical Virology (2020) 104440doi:10.1016/j.jcv.2020.104440.

[20] CDC-Centers for disease control an. prevention, Different COVID-19 vaccines, Available: https://www.cdc.gov/coronavirus/2019-ncov/vaccines/different-vaccines.html(Accessed on 20.01.2021) (15 January 2021).

[21] A. B. Gumel, E. A. Iboi, C. N. Ngonghala, E. H. Elbasha, A primer on using mathematics to understand COVID-19 dynamics. Modeling, analysis and simulations, Infectious Disease Modelling 6 148–168. doi:10.1016/j.idm.2020.11.005.

[22] A. B. Gumel, E. A. Iboi, C. N. Ngonghala, G. A. Ngwa, Mathematical assessment of the roles of vaccination and non-pharmaceutical interventions on COVID-19 dynamics: a multigroup modeling approach, medRxiv (2020). doi:10.1101/2020.12.11.20247916.

[23] L. Matrajt, J. Eaton, T. Leung, E. R. Brown, Vaccine optimization for COVID-19: who to vaccinate first?, medRxivdoi:10.1101/2020.08.14.20175257.

[24] Z. Mukandavire, F. Nyabadza, N. J. Malunguza, D. F. Cuadros, T. Shiri, G. Musuka, Quantifying early COVID-19 outbreak transmission in South Africa and exploring vaccine efficac. scenarios, PloS one 15 (7) (2020) e0236003. doi:10.1371/journal.pone.0236003.

[25] R. N. Thompson, T. D. Hollingsworth, V. Isham, D. Arribas-Bel, B. Ashby, T. Britton, P. Challenor, L. H. Chappell, H. Clapham, N. J. Cunniffe, et al., Key questions for modelling COVID-19 exi. strategies, Proceedings of the Royal Society B 287 (1932) (2020) 20201405. doi:10.1098/rspb.2020.1405.

[26] B. Tang, N. L. Bragazzi, Q. Li, S. Tang, Y. Xiao, J. Wu, An updated estimation of the risk of transmission of the novel coronavirus (2019-nCov), Infectious disease modelling 5 (2020) 248–255. doi:10.1016/j.idm.2020.02.001.

[27] A. Cori, N. M. Ferguson, C. Fraser, S. Cauchemez, A new framework and software to estimate time-varying reproduction numbers durin. epidemics, American journal of epidemiology 178 (9) (2013) 1505–1512. doi:10.1093/aje/kwt133.

[28] L. M. Bettencourt, R. M. Ribeiro, Real time bayesian estimation of the epidemic potential of emerging infectiou. diseases, PLoS One 3 (5) (2008) e2185. doi:10.1371/journal.pone.0002185.

[29] K. M. Gostic, L. McGough, E. B. Baskerville, S. Abbott, K. Joshi, C. Tedijanto, R. Kahn, R. Niehus, J. A. Hay, P. M. De Salazar, et al., Practical considerations for measuring the effective reproductive number, rt, PloS Computational Biology 16 (12) (2020) e1008409. doi:10.1101/2020.06.18.20134858.

[30] L. F. White, C. B. Moser, R. N. Thompson, M. Pagano, Statistical estimation of the reproductive number from case notificatio. data, American Journal of Epidemiology (2020). doi:10.1093/aje/kwaa211.

[31] S. R. Buckman, R. Glick, K. J. Lansing, N. Petrosky-Nadeau, L. M. Seitelman, Replicating and projecting the path of COVID-19 with a modelimplied reproductio. number, Infectious Disease Modelling (2020). doi:10.1016/j.idm.2020.08.007.

[32] M. Kiamari, G. Ramachandran, Q. Nguyen, E. Pereira, J. Holm, B. Krishnamachari, COVID-19 risk estimation using a time-varying SIR-model, in: Proceedings of the 1st ACM SIGSPATIAL International Workshop on Modeling and Understanding the Spread of COVID-19, 2020, pp. 36–42. doi:10.1145/3423459.3430759.

[33] K. Linka, M. Peirlinck, E. Kuhl, The reproduction number of COVID-19 and its correlation with public healt. interventions, Computational Mechanics 66 (2020) 1035–1050. doi:10.1007/s00466-020-01880-8.

[34] U. Abdullah, Ş. Arslan, H. S. Manap, T. Gürkan, M. Ç alışkan, A. Dayıoğlu H. N. Efe, M. Yılmaz A. Z. İbrahimoğlu, E. Gültekin, R. Durna, R. Başar F. B. Osmanoğlu, S. Ö ren, Türkiye COVID-19 pandemi izleme ekranı, Available: https://turcovid19.com/ (August 2020).

[35] Y. Demirci, Cumhurbaşkanı Erdoğan normalleşme adımlarıyla ilgili yeni kararları açıkladı., Anadolu Ajansı, Available: https://www.aa.com.tr/tr/info/infografik/18966 (Accessed on 01.11.2020) (09.06.2020).

[36] A. S. Usul, İlk CoronaVac aşı sı Sağlık Bakanı Koca’ya yapıldı., Anadolu Ajansı, Available: https://www.aa.com.tr/tr/koronavirus/ilk-coronavac-asisi-saglik-bakani-kocaya-yapildi/2108932 (Accessed on 15.01.2021) (13.01.2021).

[37] U. Abdullah, Ş. Arslan, H. S. Manap, T. Gürkan, M. Ç alışkan, A. Dayıoğlu, H. N. Efe, M. Yılmaz, A. Z. İbrahimoğlu, E. Gültekin, R. Durna, R. Başar, F. B. Osmanoğlu, S. Ö ren,Türkiye’de Covid-19 pandemisinin monitörizasyonu için interaktif ve gerçek zamanlı bir web uygulaması: TURCOVID19 (An interactive web-based dashboard for Covid-19 pandemic in real-time monitorization in Turkey: TURCOVID19), Anadolu Kliniği Tıp Bilimleri Dergisi 25 (Special Issue on COVID-19) 154–155. doi:10.21673/anadoluklin.726347.

[38] Republic of Turkey Ministery o. Health, Current status in Turkey, Available: https://covid19.saglik.gov.tr/ (January 2020).

[39] World Healt. Organization, Coronavirus disease (Covid-19) pandemic, Available: https://www.who.int/emergencies/diseases/novel-coronavirus-2019 (January 2020).

[40] European Centre for Disease Prevention and Control - An agency of the Europea. Union, Download today’s data on the geographic distribution of COVID-19 cases worldwide, Available: https://www.ecdc.europa.eu/en/publications-data/download-todays-data-geographic-distribution-covid-19-cases-worldwide (January 2020).

[41] Republic o. Turkey, Ministry of Health, Bakan Koca, TBMM’de koronavirüs ile mücadeleye ilişkin sunum yaptı., Available: https://www.saglik.gov.tr/TR,64544/bakan-koca-tbmmde-koronavirus-ile-mucadeleye-iliskin-sunum-yaptihtml. (Accessed on 01.11.2020) (19.03.2020).

[42] E. Türkmen, COVID–19 salgınında yoğun bakım ünitelerinin organizasyonu (Organization of the intensive care units during COVID-19 outbreak), Yoğun Bakım Hemşireliği Dergisi 24 (EK–1) (2020) 39–45.

[43] İrem Köker, Koronavirüs: Solunum cihazı nedir, Türkiye’de kaç adet var?, BBC Türkçe, Available: https://www.bbc.com/turkce/haberler-turkiye-52086896 (Accessed on 01.11.2020) (30.03.2020).

[44] M. Turan, AA yerli solunum cihazının üretim aşamalarını görüntüledi., Anadolu Ajansı, Available: https://www.aa.com.tr/tr/bilim-teknoloji/aa-yerli-solunum-cihazinin-uretim-asamalarini-goruntuledi/1821021 (Accessed on 01.11.2020) (28.04.2020).

[45] P. K. Bhatraju, B. J. Ghassemieh, M. Nichols, R. Kim, K. R. Jerome, A. K. Nalla, A. L. Greninger, S. Pipavath, M. M. Wurfel, L. Evans, et al., Covid–19 in critically ill patients in the Seattle region–cas. series, New England Journal of Medicine 382 (21) (2020) 2012–2022. doi:10.1056/NEJMoa2004500.

[46] Q. Bi, Y. Wu, S. Mei, C. Ye, X. Zou, Z. Zhang, X. Liu, L. Wei, S. A. Truelove, T. Zhang, et al., Epidemiology and transmission of COVID-19 in 391 cases and 1286 of their close contacts in Shenzhen, China: a retrospective cohor. study, The Lancet Infectious Diseases 20 (8) (2020) 911–919. doi:10.1016/S1473-3099(20)30287-5.

[47] N. M. Linton, T. Kobayashi, Y. Yang, K. Hayashi, A. R. Akhmetzhanov, S.-m. Jung, B. Yuan, R. Kinoshita, H. Nishiura, Incubation period and other epidemiological characteristics of 2019 novel coronavirus infections with right truncation: a statistical analysis of publicly available cas. data, Journal of clinical medicine 9 (2) (2020) 538. doi:10.3390/jcm9020538.

[48] P. Van den Driessche, J. Watmough, Reproduction numbers and subthreshold endemic equilibria for compartmental models of diseas. transmission, Mathematical biosciences 180 (1-2) (2002) 29–48. doi:10.1016/S0025-5564(02)00108-6.

[49] O. Diekman, J. Heesterbeekn, Mathematical Epidemiology of Infectious Diseases: Model Building, Analyis and Interpretation, Wiley, New York, 1999.

[50] H. Nishiura, G. Chowell, The effective reproduction number as a prelude to statistical estimation of time-dependent epidemic trends, in: Mathematical and statistical estimation approaches in epidemiology, Springer, 2009, pp. 103–121. doi:10.1007/978-90-481-2313-1_5.

[51] G. Chowell, H. Nishiura, L. M. Bettencourt, Comparative estimation of the reproduction number for pandemic influenza from daily case notificatio. data, Journal of the Royal Society Interface 4 (12) (2007) 155–166.

[52] The Worl. Bank, Countries and economies, Available: https://data.worldbank.org/country/turkey?view=chart (Accessed on 01.03.2020) (March 2020).

[53] J. D’Errico, fminsearchbnd. fminsearchcon, MATLAB Central File Exchange. (2020). URL https://www.mathworks.com/matlabcentral/fileexchange/8277-fminsearchbnd-fminsearchcon(AccessedonMarch1,2020)

[54] S. Marino, I. B. Hogue, C. J. Ray, D. E. Kirschner, A methodology for performing global uncertainty and sensitivity analysis in system. biology, Journal of theoretical biology 254 (1) (2008) 178–196. doi:10.1016/j.jtbi.2008.04.011.

[55] Centers for Diseas. Control, Estimated COVID-19 infections, symptomati. illnesses, and hospitalizations—United States, https://www.cdc.gov/coronavirus/2019-ncov/cases-updates/burden.html (Accessed on 13.01.2021) (2021).

[56] Pfizer Inc., Pfizer and Biontech conclude phase 3 study of COVID-19 vaccin. candidate, meeting all primary efficac. endpoints, Pfizer Inc., Available: https://www.pfizer.com/news/press-release/press-release-detail/pfizer-and-biontech-conclude-phase-3-study-covid-19-vaccine (Accessed on 19.11.2020) (01.12.2020).

[57] E. Mahase, Covid-19: Moderna vaccine is nearly 95% effective, trial involving high risk and elderly people shows, BMJ: British Medical Journal (Online) 371 (2020). doi:10.1136/bmj.m4471.

[58] AstraZeneca, AZD1222 vaccine met primary efficacy endpoint in preventing covid-19, AstraZeneca, Available: https://www.astrazeneca.com/content/astraz/media-centre/press-releases/2020/azd1222hlrhtml. (Accessed on 01.12.2020) (23.11.2020).

[59] G. McGregor, Why did the efficacy of China’s top vaccine drop from 78% to 50% ?, Fortune, Available: https://fortune.com/2021/01/13/sinovac-vaccine-efficacy-rate-drop/ (Accessed on 14.01.2021) (13.01.2021).

